# Large Language Models to Label CT Radiology Reports for Incidental Findings Found in Lung Cancer Screening: A Comparative Analysis

**DOI:** 10.64898/2026.11.09.26367611

**Authors:** Fennie van der Graaf, Noa Antonissen, Ernst T. Scholten, Hanke Schalkx, Mathias Prokop, Colin Jacobs

## Abstract

**Objective:** To compare LLM and prompting strategies for labeling Specific Incidental Findings (SIFs) occurring during lung cancer screening, in radiology reports.

**Materials and Methods:** In this retrospective study, 300 CT reports from a Dutch academic hospital were included between 19^th^ March 2003 and 28^th^ December 2020. GPT-5, GPT-4-Turbo, Llama-3.1-8B-INST, MedGemma-4B-IT, a keyword-based search method, and a radiologist labelled the reports for the presence or absence of nine SIFs. The radiologist’s scores were the reference standard. Each LLM was evaluated with four different prompting strategies: two without Chain-of-Thought (CoT), and two with CoT (one zero shot, one few shot). The automated labelling methods were evaluated against the radiologist primarily using Cohen’s Kappa, and additionally sensitivity, specificity, and F1 scores.

**Results:** GPT-5 achieved substantial or almost perfect agreement for seven out of nine SIFs: aortic valve disease, bronchiectasis, consolidation, coronary artery calcification, emphysema, abnormality in mediastinal lymph nodes, and thyroid abnormalities, with Cohen κ values of 0.81 (0.70, 0.93), 0.97 (0.93, 1.00), 0.71 (0.62, 0.82), 0.92 (0.87, 0.96), 0.91 (0.84, 0.97), 0.71 (0.62, 0.80), and 0.92 (0.86, 0.98) respectively. For the remaining two SIFs, moderate LLM agreement was obtained with MedGemma-4B-IT and with GPT-4-Turbo with Cohen κ values of 0.48 (0.40, 0.60), 0.42 (0.31, 0.58) for interstitial lung abnormalities and mediastinal mass respectively. The highest agreement was obtained with the non-CoT prompt strategies for most SIFs.

**Conclusion:** This study demonstrated that LLMs can automatically label radiology reports with agreement ranging from almost perfect to moderate, but not one LLM – prompt combination achieved the highest agreement across all SIFs.

## Background and Significance

In November 2022, the Council of the European Union (EU) recommended EU countries to explore the feasibility and effectiveness of Lung Cancer Screening (LCS) with use of low-dose computed tomography (LDCT).[1] Beyond the primary purpose of early detection of lung cancer, LDCT imaging may reveal a variety of incidental findings (IFs). A group of six European radiological societies published a statement about the management of IFs for LCS.[2] The statement paper named nine specific incidental findings (SIFs): aortic valve disease (AVD), bronchiectasis (Bron), consolidation (Cons), coronary artery calcification (CAC), emphysema (Emp), interstitial lung abnormalities (ILA), mediastinal mass (MM), mediastinal lymph nodes (MLN) and thyroid abnormalities (TA).

The implementation of LCS across the EU has several challenges, including the anticipated workload for radiologists to read the large volume of LDCT scans. Artificial Intelligence (AI) algorithms could support by assisting the interpretation of LDCT scans. Despite more than 15 CE-marked commercial solutions for pulmonary nodules in Europe,[3] the development of AI algorithms for SIFs is still in its infancy.

A large amount of annotated SIFs from retrospective CT scans are needed to train a SIF detection and classification algorithm. However, screening programs such as the National Lung Screening Trial (NLST) lack comprehensive annotations for SIFs,[4] they only include optional verbatim fields and a few were encoded during screening, namely: consolidation and emphysema. During the NLST, diagnoses identified during patient assessments and treatments were recorded using ICD-9 codes, however these do not link to a specific scan.[5] Clinical radiology reports can provide a rich source of annotation for SIFs and clinical CTs can enrich the training dataset. By extracting SIFs from the clinical text, radiology reports can serve as weak supervision for training models that can classify SIFs in CT scans. This process enables the creation of large labeled datasets that do not require time-intensive manual annotations.[6]

Traditional keyword search methods fail to effectively capture the nuances of medical reports, as they often miss context, cannot handle negation well and struggle with clinical terminology variations. At present, no publicly available Natural Language Processing (NLP) model exists for labelling the SIFs in LCS from CT reports, such as CheXpert/CheXbert which labels 13 findings in chest X-ray reports. Bhayana et al. 2024 demonstrated the potential of zero-shot prompting GPT-4 for identifying incidental findings in radiology reports, however there is a growing demand to validate light weight, privacy–preserving, open-source models for CT report labeling.[7] Besides model choice, prompting engineering influences LLM performance. Wei et al. 2022 introduced Chain of Thought (CoT) Prompting to encourage LLMs to think step by step to improve performance on reasoning tasks.[8] Prior research in CoT prompting in radiology by Khosravi et al. 2025 found variable effects depending on the model, and so more investigation is needed to determine if CoT is a recommended strategy in radiology.[9]

## Objective

Our aim is to investigate the best strategy for the automatic labelling of radiology reports for SIFs defined during LCS.

## Materials and Methods

This retrospective study was designed to determine the feasibility and best strategy of LLMs to annotate radiology reports for SIFs. The institutional review board approved this study and waived the need for informed consent because of the retrospective design and the use of de-identified data from an academic medical center in the Netherlands (CMO2016-3045, Project R0134021A, 2025-12-22).

### Data

For this study, we used an existing dataset consisting of all Dutch CT thorax abdomen radiology reports between 13^th^ May 1997 and 1^st^ February 2021, previously downloaded from the electronic health record (EHR) system of our institution. The reports were de-identified using an internal radiology report anonymization method that was approved by the local institutional privacy office. We applied the following exclusion criteria to the initial pool of radiology reports; patients aged less than 18, post-mortem, punction, pediatric, drainage, radiofrequency ablation (RFA), or external studies. After applying the exclusion criteria, 200 reports were randomly selected from the time period: 1^st^ of January 2016 and the 1^st^ December 2017. This dataset was then extended to increase the diversity of the dataset with 100 reports, which were randomly selected from entire date range 13^th^ May 1997 and 1^st^ February 2021. Prior to sending radiology reports to external cloud services, they were manually checked for remaining presence of personal health information, and if needed remaining personal health information was removed, according to hospital privacy regulations. A quality check was performed on the selected reports, more details can be found in Supplementary Information S1. No pre-processing of the radiology report was done, the full report was used, more details can be found in Supplementary Information S2.

### Data Annotation

The reference standard for presence of SIFs was set by a general radiologist (E.T.S.) with 32 years of experience. The radiologist was provided with the IF statement paper,[2] and was asked to label the presence or absence of the nine SIFs following the statement paper recommendations. The software used for labeling was Microsoft Excel (v2408) and Doccano (v1.8.4).[10] The following SIFs were annotated: aortic valve disease (AVD), bronchiectasis (Bron), consolidation (Cons), coronary artery calcification (CAC), emphysema (Emph), interstitial lung abnormalities (ILA), abnormalities in mediastinal lymph nodes (MLN), mediastinal mass (MM) and thyroid abnormalities (TA).

### Models

Four LLMs were evaluated in this study; two proprietary models: GPT-5,[11] and GPT-4-Turbo model,[12] via the OpenAI API, and two open-source models: Llama-3.1-8B-INST by Meta,[13] and MedGemma-4B-IT by Google.[14] As a baseline comparison, we developed a Keyword Search (regex) to also label the reports for the nine SIFs. The open-source models were deployed and run locally, further information about the labelling methods can be found in the Supplementary Information S3 and S4.

### Prompting Strategies

Four different prompting strategies were applied in this study. The first prompting strategy was a general prompt which instructed the LLM to label the report for all nine SIFs, and did not offer any detail regarding how to determine their presence or absence (Prompt V1). The second strategy divided the prompt per SIF and instructed the LLM to label the report only for the presence or absence of a single SIF, and offered additional information based on the IF Statement paper,[2] to determine the presence or absence of the SIF (Prompt V2). The third strategy was the same as the second, only it added CoT (Prompt V3). The fourth strategy was the same as the third, but few shot examples were also provided; three examples consisting each of a radiology report, reasoning, and output were provided (Prompt V4). More details on the prompting strategies can be found in the Supplementary Information S5.

### Post Processing

Post processing of the output of the models was performed to extract scores, more details can be found in Supplementary Information S6.

### Evaluation

Cohen’s Kappa was the primary metric used to assess the agreement between the automatic methods and the radiologist. The Cohen’s Kappa was interpreted into these agreement categorizations: no agreement (κ < 0), slight agreement (κ: 0.0 - 0.20), fair agreement (κ: 0.21 - 0.40), moderate agreement (κ: 0.41 - 0.60), substantial agreement (κ: 0.61 - 0.80), and almost perfect agreement (κ: 0.81 – 1.00) as suggested by Landis and Koch (1977).[15] Supporting metrics include sensitivity, specificity, and F1 score. All 95% confidence intervals were calculated using bootstrapping with 1000 bootstrap iterations. In Supplementary Information S7, more details are given regarding how the analysis into the differences between model and prompt types was performed. In Figure 1, the process of evaluation of each model – prompt combination is shown.

**Figure 1.**
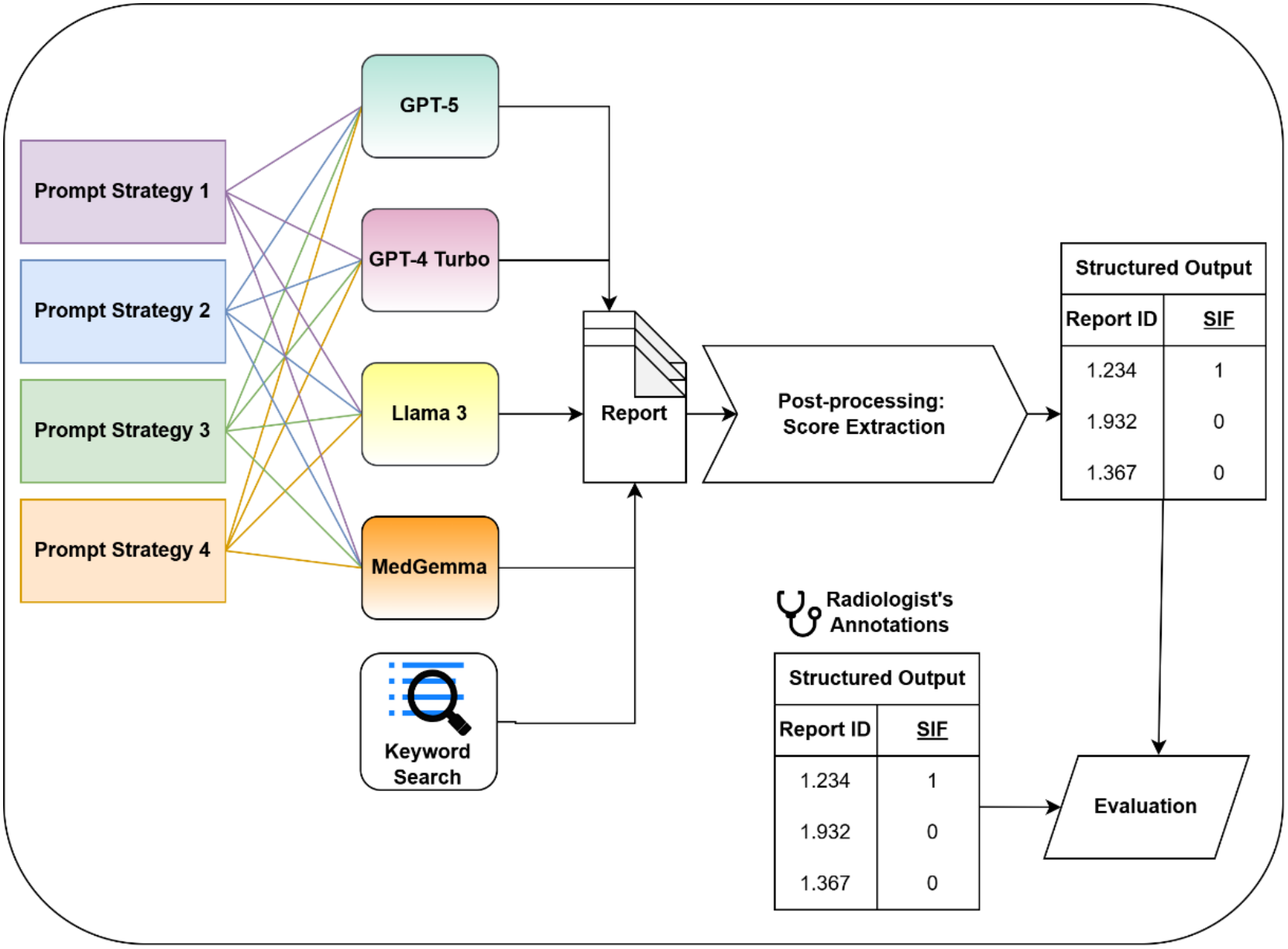
Diagram showing the workflow of the combinations of each model and prompt evaluated on each report, and then for each of these combinations the scores for the Specific Incidental Finding (SIF) were extracted from the output to generate a structured output to compare with the radiologist’s annotations. Prompt Strategies 1, 2, 3, and 4 are as follows: general prompt which instructed the LLM to label the report for all nine SIFs, prompt per SIF and provided additional information per SIF, the same as Prompt V2 only it added CoT, the same as Prompt V3 only it added few shot examples.

## Results

### Data Selection

Figure 2 shows the results of the data selection of this study. In total, we obtained 264,186 reports from the period 1997 - 2021. After applying our exclusion criteria, 212,017 reports were left for further analysis. From this set, 300 radiology reports were randomly selected. As a result of this selection, the date range of these selected 300 reports was between 19^th^ March 2003 and 28^th^ December 2020. In the Supplementary Information Table S1, the frequency of SIFs based on the radiologist’s labels are provided.

**Figure 2.**
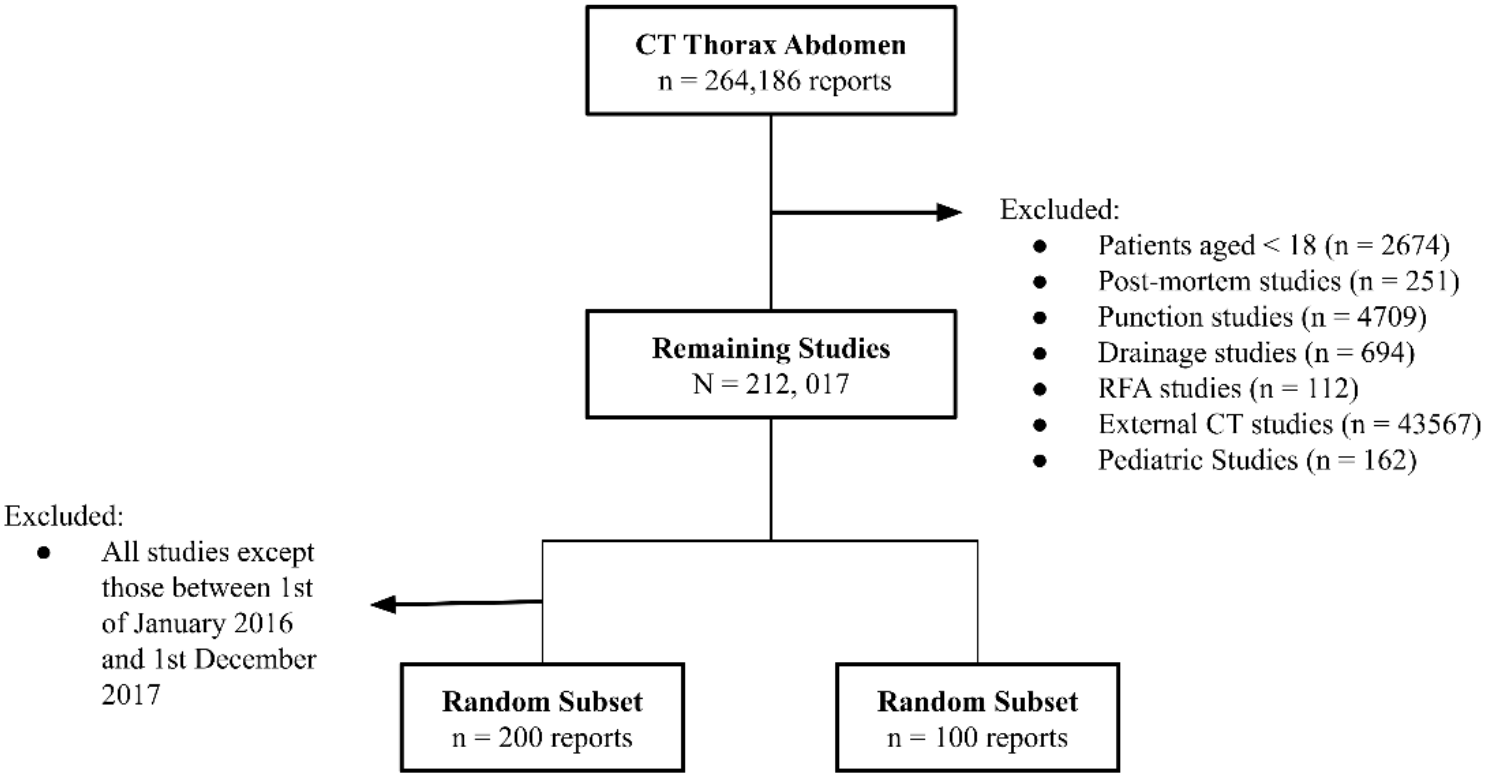
The data selection process from the internal academic hospital center and random selection of the test dataset.

### Comparing the Best Open-source and Proprietary Methods

For eight of the nine SIFs, the best proprietary LLM achieved higher agreement with the radiologist than the best open-source LLM; ILA was the exception, for which MedGemma-4B-IT achieved slightly higher agreement (κ = 0.48 [0.40, 0.60]) than the best proprietary LLM, GPT-5 (κ = 0.46 [0.37, 0.56]) (Table 1). The best proprietary and open-source LLMs had the same agreement category for five SIFs, indicating comparable levels of agreement for several findings. This was also reflected in the F1 scores in Table 2, with little difference between the best proprietary and open-source LLMs for bronchiectasis (0.97 [0.94, 1.00] vs 0.94 [0.88, 0.99]), consolidation (0.81 [0.70, 0.90] vs 0.76 [0.63, 0.87]), emphysema (0.92 [0.86, 0.97] vs 0.89 [0.81, 0.95]), and ILA (0.50 [0.34, 0.63] vs 0.52 [0.37, 0.68]), respectively. Compared with the Keyword Search, the open-source LLMs achieved higher agreement and higher F1 scores for seven of the nine SIFs; AVD and MM were the exceptions; for which Keyword Search achieved κ = 0.71 [0.48, 0.88] and κ = 0.57 [0.00, 0.89], respectively. The keyword Search resulted in lower sensitivity (0.04–0.71) (Table 3) compared with a consistently high specificity (0.94–1.00) (Table 4) across SIFs, suggesting that its lower agreement and F1 scores for most SIFs were mainly associated with missed positive findings.

**Table 1.** Pairwise Cohen’s Kappa with 95% bootstrapped confidence intervals per SIF with Highest Scores per Model Type in Bold.

| Model type | Model | Prompt | AVD | Bron | Cons | CAC | Emph | ILA | MLN | MM | TA |
|---|---|---|---|---|---|---|---|---|---|---|---|
| Proprietary | GPT-4-Turbo | V1 | 0.78 (0.63, 0.93) Substantial | 0.93 (0.87, 0.99) Almost Perfect | 0.66 (0.57, 0.75) Substantial | 0.87 (0.81, 0.93) Almost Perfect | 0.88 (0.82, 0.95) Almost Perfect | 0.39 (0.32, 0.48) Fair | 0.61 (0.53, 0.69) Substantial | **0.42 (0.31, 0.58) Moderate** | 0.89 (0.81, 0.96) Almost Perfect |
|  |  | V2 | 0.78 (0.59, 0.93) Substantial | 0.96 (0.90, 1.00) Almost Perfect | **0.78 (0.69, 0.87) Substantial** | 0.85 (0.77, 0.91) Almost Perfect | 0.87 (0.81, 0.95) Almost Perfect | 0.35 (0.28, 0.43) Fair | 0.70 (0.60, 0.79) Substantial | 0.35 (0.26, 0.49) Fair | 0.81 (0.67, 0.92) Almost Perfect |
|  |  | V3 | 0.69 (0.51, 0.86) Substantial | 0.96 (0.90, 1.00) Almost Perfect | 0.67 (0.58, 0.77) Substantial | 0.90 (0.84, 0.95) Almost Perfect | 0.86 (0.79, 0.93) Almost Perfect | 0.26 (0.21, 0.32) Fair | 0.64 (0.55, 0.74) Substantial | 0.22 (0.17, 0.30) Fair | 0.76 (0.61, 0.88) Substantial |
|  |  | V4 | 0.75 (0.61, 0.90) Substantial | 0.96 (0.90, 1.00) Almost Perfect | 0.35 (0.29, 0.41) Fair | 0.88 (0.82, 0.94) Almost Perfect | 0.85 (0.78, 0.92) Almost Perfect | 0.29 (0.22, 0.37) Fair | **0.72 (0.63, 0.80) Substantial** | 0.25 (0.11, 0.38) Fair | 0.77 (0.64, 0.89) Substantial |
|  | GPT-5 | V1 | **0.81 (0.70, 0.93) Almost Perfect** | **0.97 (0.93, 1.00) Almost Perfect** | 0.60 (0.51, 0.70) Moderate | **0.92 (0.87, 0.96) Almost Perfect** | 0.90 (0.83, 0.96) Almost Perfect | 0.37 (0.30, 0.46) Fair | 0.59 (0.50, 0.69) Moderate | 0.40 (0.30, 0.58) Fair | **0.92 (0.86, 0.98) Almost Perfect** |
|  |  | V2 | 0.74 (0.57, 0.89) Substantial | 0.85 (0.75, 0.93) Almost Perfect | 0.71 (0.62, 0.82) Substantial | 0.91 (0.85, 0.96) Almost Perfect | **0.91 (0.84, 0.97) Almost Perfect** | 0.30 (0.25, 0.37) Fair | 0.70 (0.60, 0.79) Substantial | 0.35 (0.26, 0.49) Fair | **0.92 (0.86, 0.98) Almost Perfect** |
|  |  | V3 | **0.81 (0.70, 0.93) Almost Perfect** | 0.84 (0.74, 0.93) Almost Perfect | 0.71 (0.62, 0.82) Substantial | 0.91 (0.87, 0.96) Almost Perfect | 0.90 (0.83, 0.96) Almost Perfect | 0.27 (0.22, 0.33) Fair | 0.71 (0.62, 0.80) Substantial | 0.33 (0.24, 0.46) Fair | **0.92 (0.86, 0.98) Almost Perfect** |
|  |  | V4 | **0.81 (0.70, 0.93) Almost Perfect** | 0.64 (0.49, 0.78) Substantial | 0.31 (0.25, 0.38) Fair | 0.91 (0.85, 0.96) Almost Perfect | 0.90 (0.83, 0.96) Almost Perfect | **0.46 (0.37, 0.56) Moderate** | 0.71 (0.62, 0.80) Substantial | 0.40 (0.30, 0.58) Fair | 0.91 (0.83, 0.98) Almost Perfect |
| Open-Source | Llama-3.1-8B-INST | V1 | 0.50 (0.23, 0.74) Moderate | 0.76 (0.66, 0.86) Substantial | 0.71 (0.59, 0.83) Substantial | 0.72 (0.62, 0.81) Substantial | **0.87 (0.80, 0.94) Almost Perfect** | 0.15 (0.12, 0.18) Slight | 0.49 (0.40, 0.57) Moderate | 0.22 (0.16, 0.29) Fair | 0.74 (0.61, 0.86) Substantial |
|  |  | V2 | 0.65 (0.44, 0.83) Substantial | 0.83 (0.73, 0.91) Almost Perfect | 0.47 (0.37, 0.57) Moderate | 0.77 (0.68, 0.85) Substantial | 0.86 (0.79, 0.93) Almost Perfect | 0.28 (0.23, 0.34) Fair | **0.54 (0.44, 0.65) Moderate** | 0.14 (0.11, 0.19) Slight | **0.80 (0.67, 0.90) Substantial** |
|  |  | V3 | 0.40 (0.29, 0.53) Fair | 0.80 (0.71, 0.89) Substantial | 0.39 (0.30, 0.47) Fair | 0.69 (0.60, 0.78) Substantial | 0.75 (0.67, 0.84) Substantial | 0.19 (0.14, 0.24) Slight | 0.58 (0.46, 0.69) Moderate | 0.08 (0.02, 0.12) Slight | 0.42 (0.26, 0.57) Substantial |
|  |  | V4 | 0.38 (0.22, 0.55) Fair | 0.68 (0.55, 0.81) Substantial | 0.22 (0.16, 0.27) Fair | 0.80 (0.72, 0.87) Substantial | 0.74 (0.66, 0.83) Substantial | 0.23 (0.17, 0.30) Fair | **0.54 (0.43, 0.65) Moderate** | 0.20 (0.05, 0.37) Slight | 0.56 (0.46, 0.66) Substantial |
|  | MedGemma-4B-IT | V1 | 0.59 (0.46, 0.74) Moderate | **0.93 (0.86, 0.99) Almost Perfect** | **0.73 (0.60, 0.84) Substantial** | 0.53 (0.46, 0.60) Moderate | 0.71 (0.63, 0.80) Substantial | **0.48 (0.40, 0.60) Moderate** | 0.42 (0.35, 0.50) Moderate | 0.11 (0.09, 0.14) Slight | 0.78 (0.69, 0.87) Substantial |
|  |  | V2 | 0.68 (0.55, 0.84) Substantial | 0.77 (0.68, 0.86) Substantial | 0.47 (0.39, 0.56) Moderate | **0.84 (0.78, 0.91) Almost Perfect** | 0.73 (0.64, 0.82) Substantial | 0.43 (0.33, 0.54) Moderate | 0.48 (0.39, 0.57) Moderate | 0.07 (0.06, 0.09) Slight | 0.69 (0.58, 0.81) Substantial |
|  |  | V3 | 0.38 (0.29, 0.49) Fair | 0.47 (0.40, 0.55) Moderate | 0.31 (0.25, 0.37) Fair | 0.62 (0.55, 0.69) Substantial | 0.44 (0.38, 0.52) Moderate | 0.15 (0.12, 0.19) Slight | 0.44 (0.37, 0.53) Moderate | 0.04 (0.03, 0.05) Slight | 0.29 (0.23, 0.35) Fair |
|  |  | V4 | 0.66 (0.53, 0.81) Substantial | 0.54 (0.46, 0.63) Moderate | 0.36 (0.27, 0.46) Fair | **0.84 (0.78, 0.91) Almost Perfect** | 0.36 (0.30, 0.43) Fair | 0.16 (0.13, 0.19) Slight | 0.44 (0.33, 0.54) Moderate | 0.10 (0.08, 0.13) Slight | 0.36 (0.30, 0.43) Fair |
|  | Key-Word Search | / | **0.71 (0.48, 0.88) Substantial** | 0.59 (0.44, 0.73) Moderate | 0.68 (0.53, 0.81) Substantial | 0.19 (0.08, 0.3) Slight | 0.67 (0.55, 0.79) Substantial | 0.46 (0.3, 0.63) Moderate | 0.12 (0, 0.26) Slight | **0.57 (0, 0.89) Moderate** | 0.07 (0.0, 0.19) Slight |

**Table 2.**
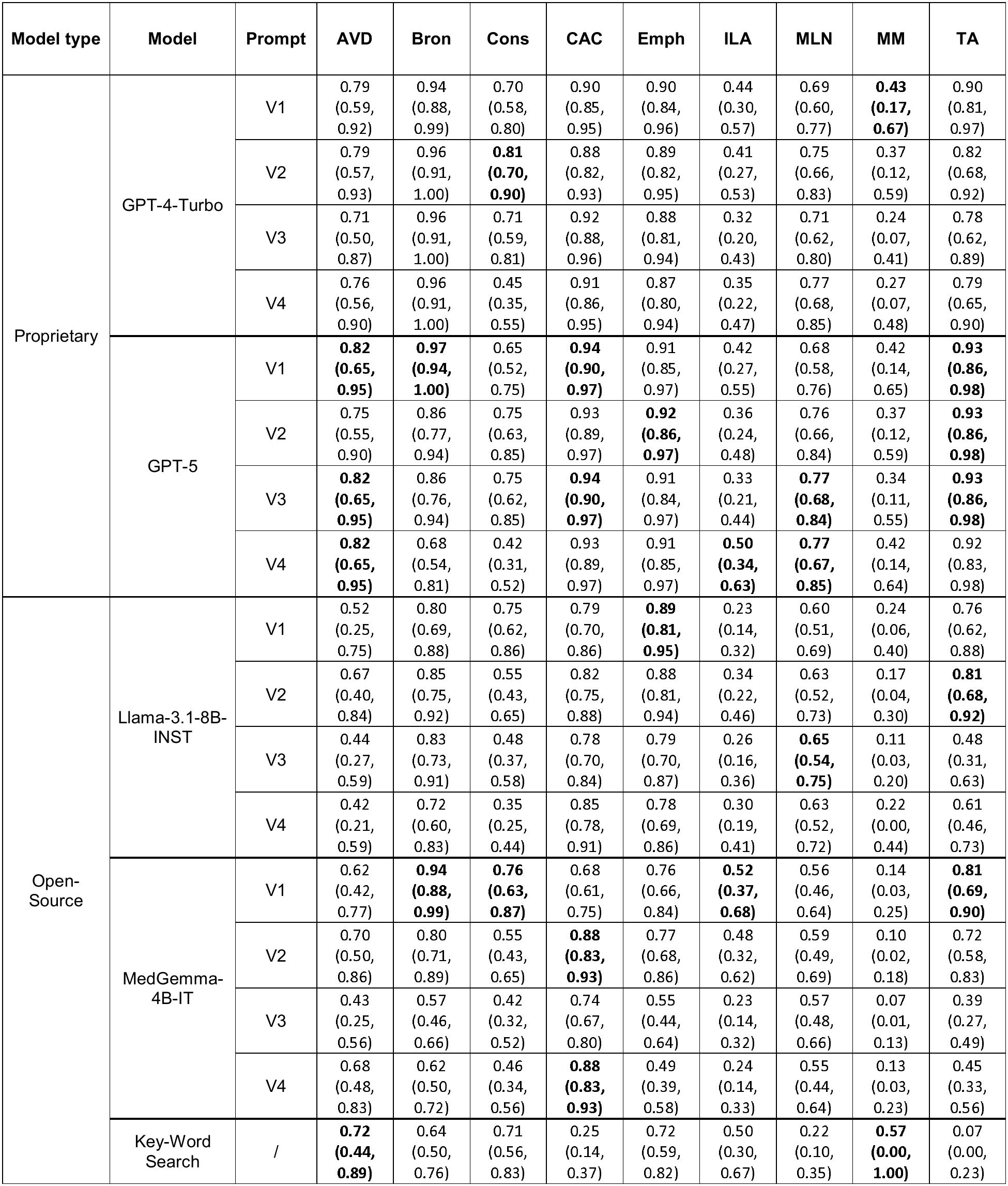
F1 Score with 95% Bootstrapped Confidence Intervals per SIF with Highest Scores per Model Type in Bold.

| Model type | Model | Prompt | AVD | Bron | Cons | CAC | Emph | ILA | MLN | MM | TA |
|---|---|---|---|---|---|---|---|---|---|---|---|
| Proprietary | GPT-4-Turbo | V1 | 0.79 (0.59, 0.92) | 0.94 (0.88, 0.99) | 0.70 (0.58, 0.80) | 0.90 (0.85, 0.95) | 0.90 (0.84, 0.96) | 0.44 (0.30, 0.57) | 0.69 (0.60, 0.77) | **0.43 (0.17, 0.67)** | 0.90 (0.81, 0.97) |
|  |  | V2 | 0.79 (0.57, 0.93) | 0.96 (0.91, 1.00) | **0.81 (0.70, 0.90)** | 0.88 (0.82, 0.93) | 0.89 (0.82, 0.95) | 0.41 (0.27, 0.53) | 0.75 (0.66, 0.83) | 0.37 (0.12, 0.59) | 0.82 (0.68, 0.92) |
|  |  | V3 | 0.71 (0.50, 0.87) | 0.96 (0.91, 1.00) | 0.71 (0.59, 0.81) | 0.92 (0.88, 0.96) | 0.88 (0.81, 0.94) | 0.32 (0.20, 0.43) | 0.71 (0.62, 0.80) | 0.24 (0.07, 0.41) | 0.78 (0.62, 0.89) |
|  |  | V4 | 0.76 (0.56, 0.90) | 0.96 (0.91, 1.00) | 0.45 (0.35, 0.55) | 0.91 (0.86, 0.95) | 0.87 (0.80, 0.94) | 0.35 (0.22, 0.47) | 0.77 (0.68, 0.85) | 0.27 (0.07, 0.48) | 0.79 (0.65, 0.90) |
|  | GPT-5 | V1 | **0.82 (0.65, 0.95)** | **0.97 (0.94, 1.00)** | 0.65 (0.52, 0.75) | **0.94 (0.90, 0.97)** | 0.91 (0.85, 0.97) | 0.42 (0.27, 0.55) | 0.68 (0.58, 0.76) | 0.42 (0.14, 0.65) | **0.93 (0.86, 0.98)** |
|  |  | V2 | 0.75 (0.55, 0.90) | 0.86 (0.77, 0.94) | 0.75 (0.63, 0.85) | 0.93 (0.89, 0.97) | **0.92 (0.86, 0.97)** | 0.36 (0.24, 0.48) | 0.76 (0.66, 0.84) | 0.37 (0.12, 0.59) | **0.93 (0.86, 0.98)** |
|  |  | V3 | **0.82 (0.65, 0.95)** | 0.86 (0.76, 0.94) | 0.75 (0.62, 0.85) | **0.94 (0.90, 0.97)** | 0.91 (0.84, 0.97) | 0.33 (0.21, 0.44) | **0.77 (0.68, 0.84)** | 0.34 (0.11, 0.55) | **0.93 (0.86, 0.98)** |
|  |  | V4 | **0.82 (0.65, 0.95)** | 0.68 (0.54, 0.81) | 0.42 (0.31, 0.52) | 0.93 (0.89, 0.97) | 0.91 (0.85, 0.97) | **0.50 (0.34, 0.63)** | **0.77 (0.67, 0.85)** | 0.42 (0.14, 0.64) | 0.92 (0.83, 0.98) |
| Open-Source | Llama-3.1-8B-INST | V1 | 0.52 (0.25, 0.75) | 0.80 (0.69, 0.88) | 0.75 (0.62, 0.86) | 0.79 (0.70, 0.86) | **0.89 (0.81, 0.95)** | 0.23 (0.14, 0.32) | 0.60 (0.51, 0.69) | 0.24 (0.06, 0.40) | 0.76 (0.62, 0.88) |
|  |  | V2 | 0.67 (0.40, 0.84) | 0.85 (0.75, 0.92) | 0.55 (0.43, 0.65) | 0.82 (0.75, 0.88) | 0.88 (0.81, 0.94) | 0.34 (0.22, 0.46) | 0.63 (0.52, 0.73) | 0.17 (0.04, 0.30) | **0.81 (0.68, 0.92)** |
|  |  | V3 | 0.44 (0.27, 0.59) | 0.83 (0.73, 0.91) | 0.48 (0.37, 0.58) | 0.78 (0.70, 0.84) | 0.79 (0.70, 0.87) | 0.26 (0.16, 0.36) | **0.65 (0.54, 0.75)** | 0.11 (0.03, 0.20) | 0.48 (0.31, 0.63) |
|  |  | V4 | 0.42 (0.21, 0.59) | 0.72 (0.60, 0.83) | 0.35 (0.25, 0.44) | 0.85 (0.78, 0.91) | 0.78 (0.69, 0.86) | 0.30 (0.19, 0.41) | 0.63 (0.52, 0.72) | 0.22 (0.00, 0.44) | 0.61 (0.46, 0.73) |
|  | MedGemma-4B-IT | V1 | 0.62 (0.42, 0.77) | **0.94 (0.88, 0.99)** | **0.76 (0.63, 0.87)** | 0.68 (0.61, 0.75) | 0.76 (0.66, 0.84) | **0.52 (0.37, 0.68)** | 0.56 (0.46, 0.64) | 0.14 (0.03, 0.25) | **0.81 (0.69, 0.90)** |
|  |  | V2 | 0.70 (0.50, 0.86) | 0.80 (0.71, 0.89) | 0.55 (0.43, 0.65) | **0.88 (0.83, 0.93)** | 0.77 (0.68, 0.86) | 0.48 (0.32, 0.62) | 0.59 (0.49, 0.69) | 0.10 (0.02, 0.18) | 0.72 (0.58, 0.83) |
|  |  | V3 | 0.43 (0.25, 0.56) | 0.57 (0.46, 0.66) | 0.42 (0.32, 0.52) | 0.74 (0.67, 0.80) | 0.55 (0.44, 0.64) | 0.23 (0.14, 0.32) | 0.57 (0.48, 0.66) | 0.07 (0.01, 0.13) | 0.39 (0.27, 0.49) |
|  |  | V4 | 0.68 (0.48, 0.83) | 0.62 (0.50, 0.72) | 0.46 (0.34, 0.56) | **0.88 (0.83, 0.93)** | 0.49 (0.39, 0.58) | 0.24 (0.14, 0.33) | 0.55 (0.44, 0.64) | 0.13 (0.03, 0.23) | 0.45 (0.33, 0.56) |
|  | Key-Word Search | / | **0.72 (0.44, 0.89)** | 0.64 (0.50, 0.76) | 0.71 (0.56, 0.83) | 0.25 (0.14, 0.37) | 0.72 (0.59, 0.82) | 0.50 (0.30, 0.67) | 0.22 (0.10, 0.35) | **0.57 (0.00, 1.00)** | 0.07 (0.00, 0.23) |

**Table 3.** Sensitivity with 95% Bootstrapped Confidence Intervals per SIF with Highest Scores per Model Type in Bold.

| **Model type** | **Model** | **Prompt** | **AVD** | **Bron** | **Cons** | **CAC** | **Emph** | **ILA** | **MLN** | **MM** | **TA** |
|---|---|---|---|---|---|---|---|---|---|---|---|
| Proprietary | GPT-4-Turbo | V1 | 0.93 (0.75, 1.00) | **1.00 (1.00, 1.00)** | **1.00 (1.00, 1.00)** | 0.97 (0.93, 1.00) | **1.00 (1.00, 1.00)** | **1.00 (1.00, 1.00)** | **0.96 (0.90, 1.00)** | **1.00 (1.00, 1.00)** | **1.00 (1.00, 1.00)** |
|  |  | V2 | 0.79 (0.54, 1.00) | 0.97 (0.91, 1.00) | **1.00 (1.00, 1.00)** | 0.88 (0.80, 0.95) | **1.00 (1.00, 1.00)** | **1.00 (1.00, 1.00)** | 0.86 (0.75, 0.95) | **1.00 (1.00, 1.00)** | 0.78 (0.60, 0.94) |
|  |  | V3 | 0.79 (0.54, 1.00) | **1.00 (1.00, 1.00)** | **1.00 (1.00, 1.00)** | 0.97 (0.93, 1.00) | **1.00 (1.00, 1.00)** | **1.00 (1.00, 1.00)** | 0.90 (0.81, 0.98) | **1.00 (1.00, 1.00)** | 0.70 (0.50, 0.88) |
|  |  | V4 | 0.93 (0.75, 1.00) | 0.97 (0.91, 1.00) | **1.00 (1.00, 1.00)** | 0.97 (0.93, 1.00) | **1.00 (1.00, 1.00)** | **0.94 (0.78, 1.00)** | **0.96 (0.90, 1.00)** | 0.80 (0.33, 1.00) | 0.78 (0.60, 0.93) |
|  | GPT-5 | V1 | **1.00 (1.00, 1.00)** | **1.00 (1.00, 1.00)** | 0.97 (0.89, 1.00) | **1.00 (1.00, 1.00)** | **1.00 (1.00, 1.00)** | **1.00 (1.00, 1.00)** | 0.90 (0.81, 0.98) | **1.00 (1.00, 1.00)** | **1.00 (1.00, 1.00)** |
|  |  | V2 | 0.86 (0.67, 1.00) | 0.82 (0.70, 0.93) | 0.97 (0.89, 1.00) | **1.00 (1.00, 1.00)** | **1.00 (1.00, 1.00)** | **1.00 (1.00, 1.00)** | 0.90 (0.81, 0.98) | **1.00 (1.00, 1.00)** | **1.00 (1.00, 1.00)** |
|  |  | V3 | **1.00 (1.00, 1.00)** | 0.79 (0.66, 0.92) | 0.97 (0.89, 1.00) | **1.00 (1.00, 1.00)** | **1.00 (1.00, 1.00)** | **1.00 (1.00, 1.00)** | 0.92 (0.84, 0.98) | **1.00 (1.00, 1.00)** | **1.00 (1.00, 1.00)** |
|  |  | V4 | **1.00 (1.00, 1.00)** | 0.54 (0.39, 0.70) | 0.97 (0.89, 1.00) | **1.00 (1.00, 1.00)** | **1.00 (1.00, 1.00)** | **1.00 (1.00, 1.00)** | 0.92 (0.84, 0.98) | **1.00 (1.00, 1.00)** | **1.00 (1.00, 1.00)** |
| Open-Source | Llama-3.1-8B-INST | V1 | 0.43 (0.18, 0.71) | 0.90 (0.79, 0.98) | 0.81 (0.67, 0.94) | 0.73 (0.62, 0.83) | 0.98 (0.92, 1.00) | **1.00 (1.00, 1.00)** | 0.92 (0.84, 0.98) | **1.00 (1.00, 1.00)** | 0.78 (0.60, 0.93) |
|  |  | V2 | 0.71 (0.44, 0.93) | 0.87 (0.76, 0.97) | 0.87 (0.74, 0.97) | 0.84 (0.75, 0.92) | **1.00 (1.00, 1.00)** | **1.00 (1.00, 1.00)** | 0.78 (0.65, 0.89) | **1.00 (1.00, 1.00)** | 0.81 (0.66, 0.96) |
|  |  | V3 | 0.86 (0.64, 1.00) | 0.92 (0.82, 1.00) | 0.90 (0.78, 1.00) | 0.85 (0.76, 0.93) | 0.98 (0.91, 1.00) | 0.94 (0.80, 1.00) | 0.74 (0.61, 0.85) | 0.80 (0.33, 1.00) | 0.56 (0.36, 0.75) |
|  |  | V4 | 0.64 (0.36, 0.90) | 0.67 (0.51, 0.82) | 0.94 (0.84, 1.00) | 0.85 (0.76, 0.93) | 0.98 (0.92, 1.00) | 0.94 (0.80, 1.00) | 0.74 (0.61, 0.86) | 0.60 (0.00, 1.00) | 0.93 (0.81, 1.00) |
|  | MedGemma-4B-IT | V1 | **0.93 (0.75, 1.00)** | 0.95 (0.87, 1.00) | 0.81 (0.66, 0.94) | **1.00 (1.00, 1.00)** | **1.00 (1.00, 1.00)** | **1.00 (1.00, 1.00)** | **0.94 (0.87, 1.00)** | **1.00 (1.00, 1.00)** | **1.00 (1.00, 1.00)** |
|  |  | V2 | **0.93 (0.75, 1.00)** | 0.95 (0.87, 1.00) | **0.97 (0.89, 1.00)** | 0.95 (0.89, 0.99) | **1.00 (1.00, 1.00)** | 0.94 (0.80, 1.00) | 0.88 (0.78, 0.96) | **1.00 (1.00, 1.00)** | 0.93 (0.81, 1.00) |
|  |  | V3 | **0.93 (0.75, 1.00)** | **0.97 (0.91, 1.00)** | **0.97 (0.89, 1.00)** | **1.00 (1.00, 1.00)** | **1.00 (1.00, 1.00)** | **1.00 (1.00, 1.00)** | **0.94 (0.86, 1.00)** | **1.00 (1.00, 1.00)** | 0.96 (0.88, 1.00) |
|  |  | V4 | **0.93 (0.75, 1.00)** | **0.97 (0.91, 1.00)** | 0.81 (0.66, 0.94) | 0.95 (0.89, 0.99) | 0.98 (0.92, 1.00) | **1.00 (1.00, 1.00)** | 0.76 (0.63, 0.88) | **1.00 (1.00, 1.00)** | **1.00 (1.00, 1.00)** |
|  | Key-Word Search | / | 0.64 (0.36, 0.89) | 0.54 (0.39, 0.69) | 0.68 (0.50, 0.84) | 0.15 (0.08, 0.23) | 0.71 (0.57, 0.85) | 0.69 (0.44, 0.90) | 0.16 (0.06, 0.28) | 0.40 (0.00, 1.00) | 0.04 (0.00, 0.13) |

**Table 4.** Specificity with 95% Bootstrapped Confidence Intervals per SIF with Highest Scores per Model Type in Bold.

| Model type | Model | Prompt | AVD | Bron | Cons | CAC | Emph | ILA | MLN | MM | TA |
|---|---|---|---|---|---|---|---|---|---|---|---|
| Proprietary | GPT-4-Turbo | V1 | 0.98 (0.96, 0.99) | 0.98 (0.96, 1.00) | 0.90 (0.87, 0.94) | 0.94 (0.91, 0.97) | **0.97 (0.94, 0.98)** | 0.86 (0.82, 0.90) | 0.84 (0.79, 0.88) | **0.96 (0.93, 0.98)** | 0.98 (0.96, 0.99) |
|  |  | V2 | **0.99 (0.98, 1.00)** | **0.99 (0.98, 1.00)** | **0.94 (0.92, 0.97)** | **0.96 (0.94, 0.99)** | 0.96 (0.94, 0.98) | 0.83 (0.79, 0.88) | **0.92 (0.88, 0.95)** | 0.94 (0.91, 0.97) | **0.99 (0.97, 1.00)** |
|  |  | V3 | 0.98 (0.96, 0.99) | **0.99 (0.97, 1.00)** | 0.91 (0.87, 0.94) | **0.96 (0.93, 0.98)** | 0.96 (0.93, 0.98) | 0.76 (0.71, 0.81) | 0.88 (0.83, 0.91) | 0.89 (0.86, 0.93) | **0.99 (0.97, 1.00)** |
|  |  | V4 | 0.98 (0.95, 0.99) | **0.99 (0.98, 1.00)** | 0.72 (0.67, 0.78) | 0.95 (0.91, 0.97) | 0.95 (0.93, 0.98) | 0.81 (0.76, 0.85) | 0.90 (0.85, 0.93) | 0.93 (0.90, 0.96) | 0.98 (0.96, 1.00) |
|  | GPT-5 | V1 | 0.98 (0.96, 0.99) | **0.99 (0.98, 1.00)** | 0.88 (0.84, 0.92) | 0.96 (0.93, 0.98) | **0.97 (0.95, 0.99)** | 0.85 (0.80, 0.88) | 0.85 (0.80, 0.89) | 0.95 (0.93, 0.98) | **0.99 (0.97, 1.00)** |
|  |  | V2 | 0.98 (0.96, 0.99) | **0.99 (0.97, 1.00)** | 0.93 (0.90, 0.96) | 0.95 (0.92, 0.98) | **0.97 (0.95, 0.99)** | 0.80 (0.76, 0.85) | 0.90 (0.86, 0.94) | 0.94 (0.92, 0.97) | **0.99 (0.97, 1.00)** |
|  |  | V3 | 0.98 (0.96, 0.99) | **0.99 (0.98, 1.00)** | 0.93 (0.90, 0.96) | 0.96 (0.93, 0.98) | **0.97 (0.95, 0.99)** | 0.77 (0.72, 0.82) | 0.90 (0.86, 0.94) | 0.94 (0.91, 0.96) | **0.99 (0.97, 1.00)** |
|  |  | V4 | 0.98 (0.96, 0.99) | **0.99 (0.98, 1.00)** | 0.70 (0.64, 0.76) | 0.95 (0.92, 0.98) | **0.97 (0.95, 0.99)** | **0.89 (0.85, 0.92)** | 0.90 (0.86, 0.94) | 0.95 (0.93, 0.98) | 0.98 (0.96, 1.00) |
| Open-Source | Llama-3.1-8B-INST | V1 | **0.99 (0.98, 1.00)** | 0.95 (0.92, 0.97) | 0.96 (0.93, 0.98) | 0.96 (0.93, 0.98) | **0.97 (0.94, 0.98)** | 0.62 (0.56, 0.69) | 0.77 (0.72, 0.82) | 0.89 (0.86, 0.93) | 0.97 (0.95, 0.99) |
|  |  | V2 | 0.98 (0.96, 0.99) | 0.97 (0.95, 0.99) | 0.85 (0.80, 0.89) | 0.94 (0.91, 0.97) | 0.96 (0.93, 0.98) | 0.78 (0.73, 0.83) | 0.86 (0.82, 0.90) | 0.83 (0.79, 0.88) | 0.98 (0.96, 1.00) |
|  |  | V3 | 0.90 (0.86, 0.93) | 0.95 (0.93, 0.98) | 0.78 (0.74, 0.84) | 0.89 (0.85, 0.93) | 0.92 (0.89, 0.95) | 0.70 (0.65, 0.76) | 0.90 (0.86, 0.93) | 0.78 (0.73, 0.83) | 0.92 (0.89, 0.95) |
|  |  | V4 | 0.93 (0.90, 0.96) | 0.97 (0.95, 0.99) | 0.60 (0.54, 0.66) | 0.95 (0.92, 0.98) | 0.92 (0.88, 0.95) | 0.76 (0.70, 0.80) | 0.88 (0.83, 0.92) | 0.94 (0.91, 0.96) | 0.89 (0.85, 0.93) |
|  | MedGemma-4B-IT | V1 | 0.95 (0.92, 0.97) | **0.99 (0.98, 1.00)** | 0.96 (0.94, 0.98) | 0.70 (0.64, 0.76) | 0.90 (0.86, 0.93) | 0.90 (0.86, 0.93) | 0.71 (0.66, 0.76) | 0.79 (0.74, 0.83) | 0.95 (0.93, 0.97) |
|  |  | V2 | 0.97 (0.94, 0.99) | 0.94 (0.91, 0.97) | 0.82 (0.77, 0.86) | 0.94 (0.91, 0.97) | 0.91 (0.87, 0.94) | 0.89 (0.85, 0.93) | 0.78 (0.73, 0.83) | 0.69 (0.64, 0.74) | 0.94 (0.91, 0.96) |
|  |  | V3 | 0.88 (0.84, 0.92) | 0.78 (0.73, 0.83) | 0.70 (0.65, 0.76) | 0.77 (0.72, 0.82) | 0.74 (0.68, 0.79) | 0.63 (0.57, 0.68) | 0.73 (0.67, 0.78) | 0.53 (0.48, 0.59) | 0.70 (0.65, 0.75) |
|  |  | V4 | 0.96 (0.94, 0.98) | 0.83 (0.78, 0.87) | 0.80 (0.75, 0.85) | 0.94 (0.90, 0.97) | 0.68 (0.63, 0.74) | 0.64 (0.58, 0.69) | 0.80 (0.75, 0.85) | 0.77 (0.72, 0.82) | 0.76 (0.71, 0.81) |
|  | Key-Word Search | / | **0.99 (0.98, 1.00)** | 0.98 (0.96, 0.99) | **0.97 (0.95, 0.99)** | **0.99 (0.97, 1.00)** | 0.96 (0.93, 0.98) | **0.94 (0.91, 0.97)** | **0.94 (0.90, 0.96)** | **1.00 (1.00, 1.00)** | **1.00 (1.00, 1.00)** |

### Comparing Model Choice

GPT-5 achieved the highest agreement among the proprietary models for six of the nine SIFs (Table 1). Overall, differences between GPT-5 and GPT-4-Turbo were small: the 95% CIs of their Cohen’s κ estimates overlapped for all nine SIFs, although the models differed in agreement category for three SIFs. The SIFs corresponding to these comparisons are provided in Supplementary Information S9. The F1 scores supported this similarity, with GPT-5 achieving a higher best F1 for the same six SIFs for which it achieved higher agreement, while GPT-4-Turbo achieved a higher F1 for consolidation and MM, both models achieved the same best F1 for MLN. Across all nine SIFs, there was little difference between their best sensitivities, specificities, and F1 scores. For example, for ILA, GPT-5 V4 and GPT-4-Turbo V1 had identical sensitivity (1.00 [1.00, 1.00]) and similar specificity (0.89 [0.85, 0.92] vs 0.86 [0.82, 0.90]), with corresponding F1 scores of 0.50 [0.34, 0.63] and 0.44 [0.30, 0.57], respectively.

MedGemma-4B-IT achieved the highest agreement among the open-source LLMs, for five of the nine SIFs (Table 1). Overall, the differences between MedGemma-4B-IT and Llama-3.1-8B-INST were greater than those of the proprietary models. The 95% CIs of the Cohen’s κ estimates for MedGemma-4B-IT and Llama-3.1-8B-INST overlapped for seven SIFs, while their agreement categories differed for four SIFs. The SIFs corresponding to these comparisons are provided in Supplementary Information S9. The F1 scores showed the same model-specific pattern: MedGemma-4B-IT achieved a higher best F1 for the same five SIFs for which it achieved higher agreement, whereas Llama-3.1-8B-INST achieved a higher F1 for emphysema, MLN, and MM, and both models achieved the same best F1 for TA. Differences in sensitivity and specificity also varied by SIF. For example, for ILA, MedGemma-4B-IT V1 and Llama-3.1-8B-INST V2 both achieved a sensitivity of 1.00 [1.00, 1.00], but MedGemma-4B-IT V1 had higher specificity (0.90 [0.86, 0.93] vs 0.78 [0.73, 0.83]), consistent with its higher F1 score (0.52 [0.37, 0.68] vs 0.34 [0.22, 0.46]), respectively.

Examples of outputted labels for the nine different SIFs from different models can be found in Figure 3.

**Figure 3.**
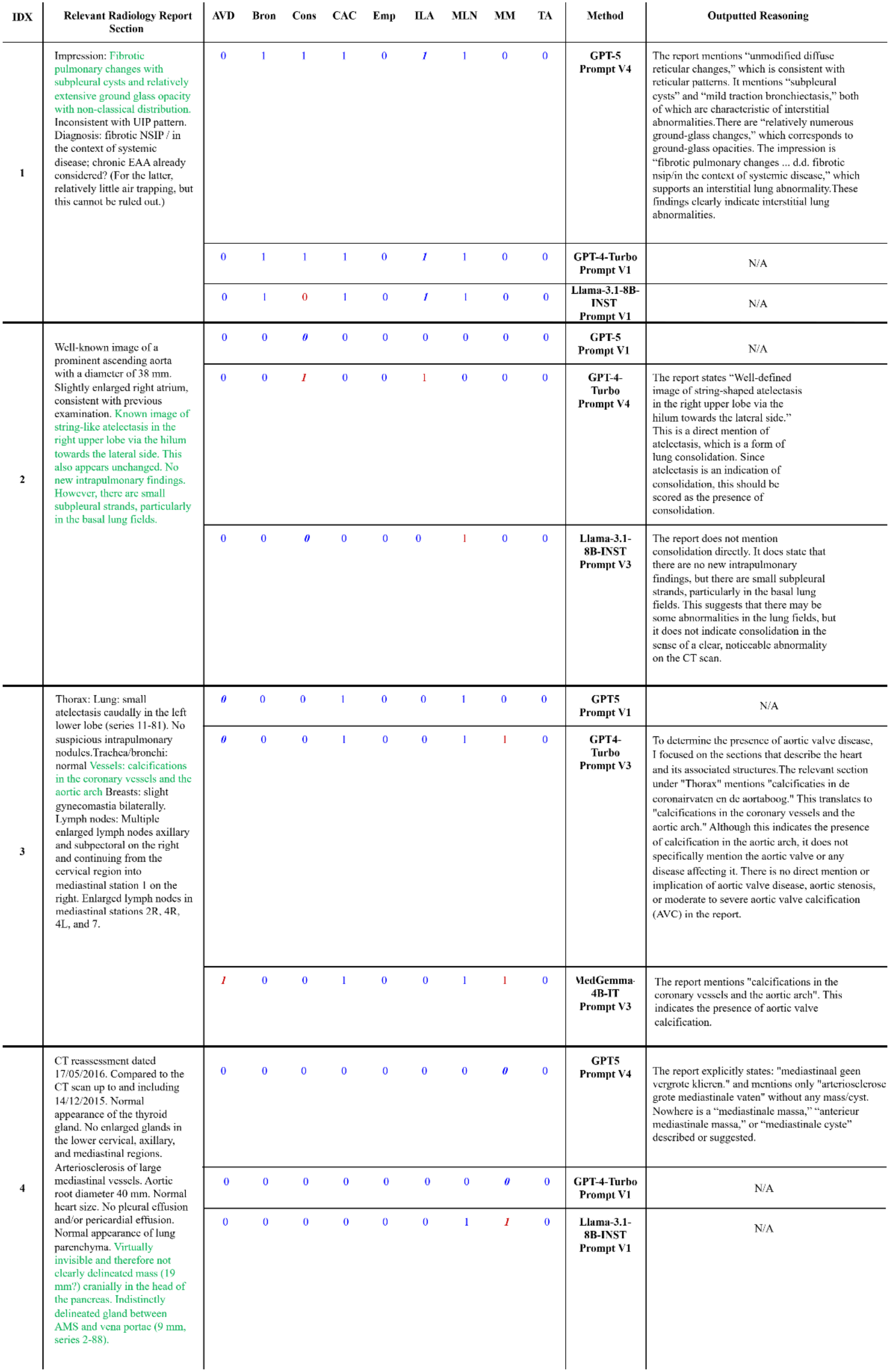
Example radiology report relevant sections and labels by the different model – prompt combinations. A value of 1 indicates presence of the SIF and a value of 0 indicates absence of the SIF. In blue the labels matched the choice of the radiologist and in red the label did not match the choice of the radiologist. The labels in bold italics refer to the SIF which was specified in the prompt strategies V2, V3, and V4. The abbreviations AVD, Bron, Cons, CAC, Emp, ILA, MLN, MM, and TA refer to: Aortic Valve Disease, Bronchiectasis, Consolidation, Coronary Artery Calcification, Emphysema, Interstitial Lung Abnormalities, Mediastinal Lymph Nodes, Mediastinal Mass, and Thyroid Abnormalities respectively.

### Comparing Prompt Choice

The majority of the highest agreement values were obtained with Prompt V1 or V2 rather than V3 or V4 for both the proprietary and open-source LLMs (Table 1). This pattern was also reflected in the F1 scores: V1 or V2 was among the highest-F1 prompts for seven of the nine SIFs for GPT-5, GPT-4-Turbo, and Llama-3.1-8B-INST, and for all nine SIFs for MedGemma-4B-IT (Table 2). The differences between prompts were more pronounced when analysing F1 scores, and less pronounced when analysing the agreement, and they varied by model and SIF.

When looking at agreement, for GPT-5 and GPT-4-Turbo, all nine SIFs had overlapping confidence intervals between the best prompt and the alternative prompts (Table 1). For each model, there was only one SIF for which the best prompt had a different agreement categorization from all three alternative prompts. For MedGemma-4B-IT and Llama-3.1-8B-INST, eight of the nine SIFs had overlapping confidence intervals between the best prompt and the alternative prompts; consolidation was the only SIF for which the confidence intervals were non-overlapping for both models (Table 1). For MedGemma-4B-IT, the best prompt had a different agreement categorization from all three alternative prompts for two SIFs, while for Llama-3.1-8B-INST this occurred for four SIFs (Table 1). The SIFs corresponding to these comparisons are provided in Supplementary Information S10.

The sensitivity, specificity, and F1 results further supported the prompt-dependent differences observed with Cohen’s κ. For example, for GPT-4-Turbo on consolidation, V2 and V4 had the same sensitivity (1.00 [1.00, 1.00]), but specificity decreased from 0.94 [0.92, 0.97] with V2 to 0.72 [0.67, 0.78] with V4. Correspondingly, F1 decreased from 0.81 [0.70, 0.90] to 0.45 [0.35, 0.55]. This indicates that the lower performance of V4 was primarily associated with an increase in falsely identified consolidation rather than missed consolidations.

In the Supplementary Information, Table S2 provides the agreement across prompt versions for each model and SIF. It shows that, across prompt versions, the open-source LLMs generally demonstrated lower agreement than the proprietary models. Examples of labels produced using the different prompting strategies are shown in Figure 3. In example report 1, GPT-5 V4 accurately justified labeling ILA as present, whereas in example report 2, GPT-4-Turbo V4 incorrectly used atelectasis as justification for labeling consolidation as present.

## Discussion

In this study, we investigated the best strategy to automatically label specific incidental findings (SIFs) occurring during Lung Cancer Screening (LCS) in CT radiology reports. This work provides insight into the first step of applying LLM-extracted pseudo-labels for training models to classify SIFs in CT scans. We compared five labelling methods: four large language models (LLMs) with four different prompting strategies and the Keyword Search method. We used Cohen’s Kappa as the primary evaluation metric. The secondary classification metrics were largely consistent with the Cohen’s κ results, with F1 reproducing the main trends observed across models and prompting strategies, while sensitivity and specificity provided complementary information on the types of classification errors.

We found that there is not a single best strategy for labelling all of the SIFs. GPT-5 performed better than all other methods for five out of nine of the SIFs. GPT-4-Turbo performed better than the other models for two of the SIFs. MedGemma-4B-IT performed better than the other models for interstitial lung abnormalities (ILA), and the Keyword Search demonstrated the best agreement for mediastinal mass (MM). For the open-source models, MedGemma-4B-IT performed better than Llama-3.1-8B-INST and the Keyword Search. However, for aortic valve disease (AVD), mediastinal mass, and interstitial lung abnormalities, the Keyword Search was either better or comparable to the open-source LLMs. In terms of prompt strategy, we observed that the prompts without Chain-of-Thought (CoT) resulted in higher agreements than the prompts with CoT. Non-CoT Prompt V1 and Prompt V2 were among the highest-performing prompting strategies for seven out of nine SIFs for GPT-5, GPT-4-Turbo, and Llama-3.1-8B-INST, and for all nine SIFs for MedGemma-4B-IT.

Lightweight open-source LLMs showed potential for labeling SIFs, since for five of the nine SIFs the agreement categorization between the best open-source LLM and the best proprietary LLM was the same. In our study, we selected lightweight open-source language models: 8B and 4B in parameter sizes. Nowak et al. 2025 found that the proprietary model GPT-4o had highest performance when extracting labels from Chest X Ray reports.[16] In their study they compared both light weight (8B) and larger open-source models to GPT-4o and found that the larger open-source models such as Mistral-Large (123B) had comparable performance with GPT-4o. The impressive performance of lightweight MedGemma-4B-IT in our study was likely based on its extensive fine-tuning on radiological images and their associated reports. Zhang et al. 2024 demonstrated that fine-tuning of an open-source LLM can result in comparable performance with GPT-4 for medical evidence summarization.[17] Furthermore, other studies also found that LLMs perform better than rule-based methods for label extraction.[18] However in our study, the Keyword Search performed best for labelling mediastinal mass. For mediastinal mass there was low linguistic variability, and the LLMs tended to label reports which mentioned masses anywhere (not specifically in the mediastinal region) or tended to label enlargement of lymph nodes as mediastinal mass. Similarly, Mukerjee et al. 2023 reported that for some classes, the rule-based system outperformed Vicuna.[18]

There are several advantages for using lighter weight, open-source models for medical applications. Savage at al. 2025 outlined general advantages, such as customizability, flexibility for commercial use, and security – as open-source LLMs can be fully downloaded to run locally on-site or in institutionally managed cloud instances.[19] In terms of size, running smaller models requires less resources to run and this translates to lower cost, greater portability, and lower environmental impact. In this study, we have demonstrated that for labelling some of the SIFs, lighter weight open-source methods offered better, or comparable performance to proprietary, heavier models. However, we found that the light-weight open-source models were more affected by prompt engineering than the proprietary models. Smaller models may be more affected by prompt engineering because they have less representational capacity (lower capabilities to fill in missing context from the prompt) and they are fine-tuned on small instruction datasets with limited prompt diversity.

Our results demonstrated that CoT prompting yielded lower agreement with the radiologist. Khosravi et al. 2025 also found that CoT led to poorer results for smaller models for determining the presence or absence of a finding in radiology reports.[9] They hypothesized that due to model ‘overthinking’, performance on simple tasks is reduced, and concluded that prompting strategy should be task specific. For both the open-source and the proprietary models we found that sometimes the reasoning output for a label was nonsensical. For example, GPT-4-Turbo/Prompt V4 wrongly justified labelling presence for consolidation because the report mentioned atelectasis. In many reports consolidation and atelectasis are often named together and pre-training on such reports may have resulted in this misunderstanding. Another example, MedGemma-4B-IT/Prompt V3 incorrectly labelled for the presence of AVD when the report only referenced the aortic arch.

In this work, we did not find an improvement in agreement when few shot examples were provided in the CoT (Prompt V4). This result was similarly found by Nowak et al. 2025, and they speculated that examples led some LLMs to allocate less attention to the instruction part of the prompt.[16] In longer prompts, LLMs tend to focus on information at the beginning and end of the instructions, and that performance reduces when models must access relevant information in the middle of prompts.[20] This may suggest that CoT is better suited for tasks with shorter examples, rather than for our task which required the entire radiology report as an example.

There are several limitations to this study. Firstly, the size of our dataset (300 reports) resulted in only five examples of mediastinal mass. While in a screening setting the prevalence of mediastinal mass would be very low, this small number reduces the certainty of our conclusions regarding mediastinal mass. Given that it took the radiologist about two hours to label 100 reports, for feasibility purposes 300 reports were selected. An extensive analysis using a larger corpus of reports would help future studies to guarantee a substantial number of cases for each SIF. Secondly, a single radiologist served as the reference standard. As inter-reader agreement was not assessed, we cannot determine whether disagreement between the models and the reference labels reflects label noise, model limitations or intrinsic reader variability. Future work should include multiple readers and report human-human agreement. Thirdly, we investigated only the labelling of Dutch radiology reports from a single center, therefore the generalizability of the results to other centers with other languages should be investigated in future studies.

Additionally, as this was an exploratory investigation which evaluated the agreement between the models and a radiologist, the statistical significance of differences between models was not computed. Furthermore, the prompt experiment of few shot learning without CoT was not included in this study, limiting our conclusions regarding the influence of few shot learning. In this study, a variety of LLMs in terms of model size, proprietary status, release date are used.

We acknowledge that this field is rapidly evolving and that these evaluations are inherently time dependent, and may not fully reflect the performance of future LLMs.

In conclusion, this study evaluated the best strategy of using large language models to automatically label lung cancer screening–relevant incidental findings in Dutch radiology reports. LLMs reached moderate or above agreement with a radiologist for labelling radiology reports for SIFs. For five out of the nine SIFs, open-source lighter LLMs had comparable or higher agreement with a radiologist than proprietary, heavier LLMs. There was not one optimal model/prompt strategy across SIFs, and CoT-prompting reduced performance. GPT-5 reached almost perfect agreement for most SIFs, while MedGemma-4B-IT demonstrated substantial or above performance for most SIFs. These results support using LLM-based report labeling as a weak-annotation approach to help scale training data for future incidental finding algorithms. For future weakly supervised labeling pipelines, we suggest starting with concise non-CoT prompts and if privacy is essential: lightweight open-source models. We recommend selecting the optimal model and prompt separately for each finding using an expert labelled validation dataset, before being deployed to generate large-scale pseudo-labels.

## Data Availability

The dataset used in this research is not publicly available.

## Acknowledgements

We would like to thank Luc Builtjes and Lisa Klok for their support when reviewing and editing this work. This study is part of the project ROBUST: Trustworthy AI-based Systems for Sustainable Growth with project number KICH3.L TP.20.006, and has received funding from the Dutch Science Foundation, the Dutch Ministry of Economic Affairs, and MeVis Medical Solutions (Bremen, Germany).

