## Supplementary Material for "Large Language Models to Label CT Radiology Reports for Incidental Findings Found in Lung Cancer Screening: A Comparative Analysis"

### Supplementary Information:

**Part 1: Methods**

1. **Reports Quality Check**

These were manually checked for good report quality (at least two sentences describing the scan) and three reports were found to be of poor quality, but were still included in the analysis.

2. **Pre-processing of Reports**

There was no pre-processing performed. Similar performance was observed with a small sample when translating the reports to English vs keeping them in Dutch, so the reports were kept in Dutch.

3. **LLM Inference**
1. GPT-5: Inference run via OpenAI API with openai v1.99.9 with a set random torch seed, temperature could not be user-set since it is a reasoning model, the reasoning parameter used was the default parameter (medium), the number of completions per request was set n = 1, the max completion token number was set to 3000, and the presence penalty was set to 0.0 since scoring was requested in a strict format.
2. GPT-4-Turbo: Inference run via OpenAI API with openai v1.75.0 , with a set random torch seed, temperature=0.2 since some randomness in generation is desired for the CoT prompts, the number of completions per request was set n=1, the max completion token number was set to 1000, and the presence penalty was set to 0.0 since scoring was requested in a strict format.
3. Llama-3.1-8B-INST: This model was accessed from Huggingface: <https://huggingface.co/meta-llama/Llama-3.1-8B-Instruct>

Inference was run locally with 11GB RTX 2080 TI GPU, with a set random torch seed, 4-bit-quantization, bfloat16, max new tokens set to 1000, eos token ids <|eot_id|> applied, temperature set to 0.2, and top_p set to default value 0.9.

4. MedGemma-4b-IT: This model was accessed from Huggingface: <https://huggingface.co/google/medgemma-4b-it>

Inference run locally with 11GB RTX 2080 TI GPU, with a set random torch seed, max new tokens set to 1000, temperature set to 0.2, and top_p set to default value 0.9.

4. **Keyword Search**

In Dutch radiology reports, the reports can be written in a combination of English terms and Dutch terms. The following terms (case insensitive) per SIF were searched for in report texts.

1. Interstitial lung abnormalities: 'interstitial lung abnormalities', 'interstitiele longafwijkingen', 'fibrose', 'ground glass opacities', 'tractie bronchiectasieen'
2. Emphysema: 'emphysema', 'emfyseem', 'emfysemateuze verandering'
3. Bronchiectasis: 'bronchiectasis', 'bronchiectasie', 'bronchiectasieen'
4. Consolidation: 'consolidatie', 'infiltraat'
5. Coronary calcification": ‘coronary calcification’, ‘coronairsclerose’, ‘coronair atherosclerose’, ‘coronairkalk’
6. Aortic valve disease: 'aortic valve disease', 'aortaklepaandoening', 'aortaklepsclerose', 'aortaklepverkalking', 'aortaklep kalk', 'aortaklep'
7. Mediastinal mass: 'mediastinal mass', 'mediastinale massa'
8. Mediastinal lymph nodes: 'mediastinal lymph nodes', 'mediastinale lymfeklieren',

'klieren mediastinaal', 'lymfklieren mediastinaal'

9. Thyroid abnormalities: 'thyroid abnormalities', 'schildklierafwijkingen',

'schildklier nodule', 'schildklier nodus'

The following negations were applied before each terms, and if the negation was found with a term it was counted as an absence of the SIF:

not|no|geen|niet|nooit|zonder|never

5. **Prompting**

The system prompt was written in English to use the original English terms indicated in the statement paper for the incidental findings (2), and since the LLMs are pre-dominantly trained with English data, better performance was observed with an English system prompt.

**Prompt V1**

You are an expert Dutch radiologist. You are qualified to review and process Dutch medical records of patients. You specialize in reading Chest CT; you understand the anatomy of the lungs. Your job is to determine if the subject is interstitial lung abnormalities, emphysema**,** bronchiectasis, consolidation, coronary artery calcification, aortic valve disease, mediastinal mass, mediastinal lymph nodes, thyroid abnormalities in the text.

If the topic is present, give a score of 1; if it is not present, give a score of 0. More than one topic can be given a score 1 or a score of 0.

It is possible that none of the topics are present in the text, so then give each topic a score 0.

IMPORTANT: Do not create your own topics, only score the topics I reported:

interstitial lung abnormalities, emphysema, bronchiectasis, consolidation, coronary artery calcification, aortic valve disease, mediastinal mass, mediastinal lymph nodes, or thyroid abnormalities appears in the text.

IMPORTANT: You should score based on diagnoses mentioned in the text as well as diagnoses suggested by the text.

IMPORTANT: Report the score in an exact, concise list, with little additional explanation. Use the following format for each diagnosis:'subject: score'. Make sure the format is followed exactly.

IMPORTANT: The score can only be 0 or 1.

**Prompt V2 (Example: ILA)**

You are an expert Dutch radiologist. You are qualified to review and process Dutch medical records of patients. You specialize in reading Chest CT reports. Your job is to determine if in the given report there is mention of substantial interstitial lung abnormalities.

If interstitial lung abnormalities are present, give a score of 1; if it is not present, give a score of 0.

IMPORTANT: Do not create your own topics, only score the presence or absence of interstitial lung abnormalities.

IMPORTANT: You should score based on direct mention of interstitial lung abnormalities in the text and based on signs that interstitial lung abnormalities is suggested by the text. Mention of the following should also be scored as interstitial lung abnormalities:

- Non-dependent lung abnormalities including reticular patterns, traction bronchiectasis, non-emphysematous cyst, and ground glass opacities.

- Characterization of ILA as non-subpleural, subpleural non-fibrotic, or subpleural fibrotic.

IMPORTANT: Report the score in an exact, concise list, with little additional explanation. Use the following format 'interstitial lung abnormalities: score'. Make sure the format is followed exactly.

IMPORTANT: Report the score as only 0 or 1.

**Prompt V3 (Example: AVD)**

You are an expert Dutch radiologist. You are qualified to review and process Dutch medical records of patients. You specialize in reading Chest CT reports. Your job is to determine if in the given report there is mention of aortic valve disease.

If aortic valve disease is present, give a score of 1; if it is not present, give a score of 0.

IMPORTANT: Do not create your own topics, only score the presence or absence of aortic valve disease.

IMPORTANT: You should score based on direct mention of aortic valve disease in the text and based on signs that aortic valve disease is suggested by the text. Mention of the following should also be scored as aortic valve disease:

- aortic valve disease severity correlated with aortic stenosis

- moderate or severe aortic valve calcification (AVC)

IMPORTANT: Output the reasoning and score once only. Do not repeat your output.

IMPORTANT: Do not create your own examples, score only the following report.

**Chain‑of‑Thought Instructions**

Before you state your final answer, reason step by step if aortic valve disease is mentioned in the report. In your reasoning, quote the exact phrases from the report which you use to make your decision.

IMPORTANT: Follow this output format exactly.

**Output Format**

------

\### REASONING###

…your train of thought here…

\### SCORE ###

aortic valve disease : <0|1>

Now analyse the following report:

**Prompt V4 (Example: bronchiectasis), note the actual radiology report examples were selected from the 212, 017 but not overlapping with the 300 test set. These reports were labelled by the radiologist and the reasoning was added under the supervision of the radiologist. Due to privacy restrictions, the full report cannot be included in this publication, they have been instead replaced with fake texts generated by ChatGPT.**

You are an expert Dutch radiologist. You are qualified to review and process Dutch medical records of patients. You specialize in reading Chest CT reports. Your job is to determine if in the given report there is mention of moderate or severe bronchiectasis.

If bronchiectasis is present, give a score of 1; if it is not present, give a score of 0.

IMPORTANT: Do not create your own topics, only score the presence or absence of bronchiectasis.

IMPORTANT: You should score based on direct mention of bronchiectasis in the text and based on signs that bronchiectasis is suggested by the text. Mention of the following should also be scored as bronchiectasis:

- airways larger than the adjacent pulmonary artery

- lack of tapering

- airway diameter two or more times that of the adjacent artery

**Chain‑of‑Thought Instructions**

Before you state your final answer, reason step by step if bronchiectasis is mentioned in the report. In your reasoning, quote the exact phrases from the report which you use to make your decision.

Example 1:

Verslag:

Op de CT-thorax worden milde bronchiectatische veranderingen gezien in de middenkwab en lateraal in de rechter onderkwab. Er is geen sprake van consolidaties of verdachte nodulaire afwijkingen. Het longparenchym toont verder een normale verdeling zonder aanwijzingen voor interstitiële afwijkingen.

Impressie:

Verspreid perifere bronchiectasieën, met name in de middenkwab en rechter onderkwab. Geen aanwijzingen voor acute infectie of maligniteit.

**Output**

\### REASONING###

Het rapport vermeldt "milde bronchiectatische veranderingen in de middenkwab en lateraal in de rechter onderkwab", dus bronchiëctasieën worden genoemd, maar mild, niet middelmatig of ernstig.

Maar het rapport vermeldt in de impressie "verspreid perifere bronchiectasieen", dus omdat het diffuus is en het in de impressie wordt genoemd, is het waarschijnlijk substantieel.

\### SCORE ###

bronchiectasis : 1

Example 2:

Verslag:

Op de CT-thorax worden milde bronchuswandverdikkingen gezien. Daarnaast zijn er tractiebronchiëctasieën zichtbaar in het kader van interstitiële longziekte (ILD). Er is geen sprake van consolidaties of verdachte nodulaire afwijkingen.

Impressie:

Tractiebronchiëctasieën passend bij ILD. Geen primaire bronchiectasieën of tekenen van acute infectie.

**Output**

\### REASONING###

Het rapport vermeldt "tractiebronchiectasieen" in het kader van ILD, geen primaire bronchiectasien.

Het rapport vermeldt "bronchuswand verdikking", maar geen bronchiectasie of verwijding van bronchus.

\### SCORE ###

bronchiectasis : 0

Example 3:

Verslag:

Het hart is normaal van omvang zonder aanwijzingen voor pericardvocht. De centrale en perifere lymfeklieren zijn niet vergroot. Daarnaast zijn er subtiele aanwijzingen voor kleine luchtwegziekte zichtbaar in de perifere longvelden.

Impressie:

Kleine luchtwegziekte. Geen cardiomegalie of lymfadenopathie.

**Output**

\### REASONING###

Het rapport vermeldt "kleine luchtweg ziekte", maar dit is niet hetzelfde als bronciectase omdat het vaak omkeerbaar is, terwijl bronciectase een chronische aandoening is die niet omkeerbaar is.

\### SCORE ###

bronchiectasis : 0

Now analyse the following report:

6. **Post-processing of the Labeling Raw Outputs**

The raw output from the LLM for Prompt V1 was in the format Topic: (IF), Score: <0|1>, for Prompt V2 the SIF was specified in the prompt, so for example; Coronary Artery Calcification: 0. Prompt V3 and V4 there was Reasoning: (reasoning), and then the score. From the outputted text, the score was extracted using Regex (the first value after ‘Score’, or after ‘Reasoning’ and then ‘Score’. The first value was selected since for some LLMs such as MedGemma-4B-IT, it would provide the score for the prompted report, but it would output a similar format as with Prompt V4; it would output its own examples afterwards. Additionally, LLMs would sometimes write terms a bit differently making extraction from Regex patterns challenging; such as replying with the Dutch term for the SIF rather than the English one; ‘Emfyseem: 0’ instead of ‘Emphyema: 0’. Hence if the Regex could not automatically extract the score, then it was manually extracted. There were occasions, particularly with the poor quality reports that some LLMs (MedGemma-4B-IT) would not output a score. For one report it outputted that the report was not processable, for other reports it provided a reasoning which explained that the SIF was not present, but did not directly offer a score. In the case that a clear ‘1’ for the presence of a SIF, could not be extracted, the LLM was re-run a maximum of 2x over the select reports, which for the majority of these cases did result in an outputted score. For the few that still did not result in an outputted score, a 0 was manually attributed for that output. In total, out of 8400 runs (28 experiments x 300 reports), there were 36 reports which outputted one of the formatting issues listed above; meaning that out of all runs of the LLMs, 0.43% of them required manual intervention in the post processing.

7. **Analysis of the difference between the highest scoring proprietary or open-source model/prompt combination and the alternative model’s scores, and analysis of the difference between the best prompt version and the alternative prompt versions.**

The criteria to consider a Kappa score different for a particular SIF between the highest model/prompt and alternative models was to consider whether confidence intervals were overlapping, and whether agreement categorizations were different. To consider that there was non-overlapping confidence intervals for a SIF, all four Cohen κ values from the alternative model’s prompts had to have confidence intervals that were non-overlapping with those of the highest model/prompt strategy, otherwise that SIF is considered to have overlapping confidence intervals. To consider that a SIF had a differing agreement categorization, the alternative model with all four prompt strategies should have different agreement categorizations compared to the highest model/prompt strategy. The same above criteria applied when analysing whether there was a difference between the best prompt and the alternative prompts. To consider confidence intervals as non-overlapping for a SIF, all three alternative prompt versions needed to have non-overlapping confidence intervals with the best prompt version. To consider an agreement categorization as different for a SIF, all three alternative prompt versions needed to have a different agreement categorization compared to the best prompt version.

**Part 2: Results**

8. **Frequency of SIFs in Test Dataset**

In Table S1, the frequency of SIFs that were labelled in the test set of 300 reports labelled by the radiologist are shown. There were 128 reports without any SIF label.

**Table S1. Frequency of SIFs labelled within the test dataset (n = 300) by the radiologist**

| **SIF** | **Frequency** |
| --- | --- |
| **Aortic valve disease** | 14 |
| **Bronchiectasis** | 39 |
| **Consolidation** | 31 |
| **Coronary artery calcification** | 73 |
| **Emphysema** | 41 |
| **Interstitial lung abnormalities** | 16 |
| **Mediastinal lymph nodes** | 50 |
| **Mediastinal mass** | 5 |
| **Thyroid abnormalities** | 27 |

9. **The Difference Between the Best Model/Prompt per Proprietary and Open-source LLMs and the Alternative Model Versions**

Here we provide extended analysis from Section Model Choice Effect on Agreement, regarding the difference between the best model/prompt version and the alternative model’s options (Table 1, main text). Starting with the proprietary models: in terms of confidence intervals, we considered that the best prompt/model strategy for all SIFs had overlapping confidence intervals with the alternative model/prompt strategies and hence little difference was observed between GPT-5 and GPT-4-Turbo (Table 2 main text). The best strategy for AVD: GPT-5/Prompt V1/V3/V4 had overlapping confidence intervals with all GPT-4-Turbo alternative prompts. The best strategy for bronchiectasis, GPT-5/Prompt V1 had overlapping confidence intervals with all GPT-4-Turbo alternative prompts. GPT-4-Turbo/Prompt V2 was the best strategy for consolidation, and had overlapping confidence intervals with three out of four GPT-5 alternative prompts. For CAC, the best strategy was GPT-5/Prompt V1 and it had overlapping confidence intervals with all GPT-4-Turbo alternative prompts. For emphysema, the best strategy was GPT-5/Prompt V2, which had overlapping confidence intervals with all GPT-4-Turbo alternative prompts. For ILA, the best strategy was GPT-5/Prompt V4, and had overlapping confidence intervals with three out of four GPT-4-Turbo alternative prompts. For MLN, the best strategy was GPT-4-Turbo/Prompt V4, and it had overlapping confidence intervals with all GPT-5 alternative prompts. For MM, the best strategy was GPT-4-Turbo/Prompt V1, which had overlapping confidence intervals with all GPT-5 alternative prompts. For TA, the best strategies were GPT-5/Prompt V1/V2/V3 and they had overlapping confidence intervals with all GPT-4-Turbo alternative prompts. We can observe differences in agreement categories for three SIFs between GPT-5 and GPT-4-Turbo (Table 2 main text). GPT-5/Prompt V1/V3/V4 had almost perfect agreement (0.81 [0.70, 0.93]), whilst all other GPT-4-Turbo prompt combinations had substantial agreement for AVD. Additionally, GPT-5/Prompt V4 had moderate agreement, whilst all other GPT-4-Turbo prompt combinations had fair agreement for ILA. Furthermore, GPT-4-Turbo/Prompt V1 had moderate agreement (0.42 [0.31, 0.58]), while all other GPT-5 prompt combinations had fair agreement for MM.

Following with the open-source LLMs, we found that across the SIFs there were slight differences in agreement between MedGemma-4B-IT and Llama-3.1-8B-INST. Their Cohen κ values had overlapping confidence intervals for seven SIFs. For AVD, the best strategy was MedGemma-4B-IT/Prompt V2 and it had overlapping confidence intervals with three out of four alternative Llama-3.1-8B-INST versions. For bronchiectasis, the best strategy was MedGemma-4B-IT/Prompt V1 and it had overlapping confidence intervals with three out of four alternative Llama-3.1-8B-INST versions. For consolidation, the best strategy was MedGemma-4B-IT/Prompt V1 and it had overlapping confidence intervals with one out of four Llama-3.1-8B-INST versions. For CAC, the best strategy was MedGemma-4B-IT Prompt V2/V4 and it had overlapping confidence intervals with all alternative Llama-3.1-8B-INST versions. For emphysema, the best strategy was Llama-3.1-8B-INST/Prompt V1 and it had overlapping confidence intervals with two alternative MedGemma-4B-IT versions. For ILA, the best strategy was MedGemma-4B-IT/Prompt V1 and it did not have overlapping confidence intervals with any of the four alternative Llama-3.1-8B-INST versions. For MLN, the best strategy was Llama-3.1-8B-INST/Prompt V3 and it had overlapping confidence intervals with all alternative MedGemma-4B-IT versions. For MM, the best strategy was Llama-3.1-8B-INST/Prompt V1 and it did not have overlapping confidence intervals with any of the four alternative MedGemma-4B-IT versions. For TA, the best strategy was Llama-3.1-8B-INST/Prompt V2 and it had overlapping confidence intervals with two alternative MedGemma-4B-IT versions. We can observe differences in agreement categories for four SIFs between MedGemma-4B-IT and Llama-3.1-8B-INST (Table 2 main text). MedGemma-4B-IT/V2/V4 had almost perfect agreement (0.84 [0.78, 0.91]) whilst the alternative Llama-3.1-8B-INST prompt combinations had substantial agreement for CAC. Additionally, MedGemma-4B-IT/Prompt V1 had moderate agreement (0.48 [0.40, 0.60]) whilst the alternative Llama-3.1-8B-INST prompt combinations had fair or slight agreement for ILA. Llama-3.1-8B-INST/Prompt V1 had almost perfect agreement (0.87 [0.80, 0.94]) whilst the alternative MedGemma-4B-IT prompt combinations had substantial, moderate, and fair agreement for emphysema. Additionally, Llama-3.1-8B-INST/Prompt V1 had fair agreement (0.22 [0.16, 0.29]) whilst the alternative MedGemma-4B-IT prompt combinations had slight agreement for mediastinal mass.

10. **How Different the Best Prompt Version was Compared to the Alternative Prompt Versions for that Model**

Here we provide extended analysis from Section Prompting Effect on Agreement, regarding the difference between the best prompts and the alternative prompt options (Table 2, main text). For the proprietary models, all SIFs were considered to have overlapping confidence intervals. For GPT-4-Turbo, for AVD, bronchiectasis, CAC, emphysema, ILA, MLN, and TA, all the confidence intervals of the alternative prompt versions overlapped with those of the best prompt. For consolidation and MM, one prompt version had confidence intervals which did not overlap with the best prompt version. For GPT-5, for AVD, CAC, emphysema, MLN, MM, and TA, all the confidence intervals of the alternative prompt versions overlapped with those of the best prompt. For bronchiectasis, consolidation, and ILA, one prompt version had confidence intervals which did not overlap with the best prompt version. Each proprietary model had one SIF for which the agreement categorization was considered different. GPT-4-Turbo/Prompt V1 had moderate agreement (0.42 [0.31, 0.58]), whilst the alternative prompts resulted in fair agreement for mediastinal mass. GPT-5/Prompt V4 resulted in moderate agreement for ILA whilst the alternative prompts resulted in fair agreement.

For MedGemma-4B-IT, eight of the nine SIFs were considered to have overlapping confidence intervals; consolidation was the only SIF for which the best prompt did not have overlapping confidence intervals with any of the three alternative prompts. For AVD, there was one non-overlapping prompt version with the best prompt. For bronchiectasis, CAC, emphysema, ILA, and TA, two out of three alternative prompts did not have overlapping confidence intervals with the best prompt version. For consolidation, all three alternative prompts had non-overlapping confidence intervals with the best prompt version. For MLN, all alternative prompt versions had overlapping confidence intervals with the best prompt version, while for MM one out of three alternative prompts had non-overlapping confidence intervals with the best prompt version. MedGemma-4B-IT was considered to have different agreement categorizations for two SIFs. MedGemma-4B-IT/Prompt V1 had almost perfect agreement (0.93 [0.86, 0.99]) for bronchiectasis, whilst the other prompts resulted in substantial or moderate agreement. Additionally, MedGemma-4B-IT/Prompt V1 had substantial agreement (0.73 [0.60, 0.84]) whilst the other prompts had moderate or fair agreement for consolidation.

For Llama-3.1-8B-INST, eight of the nine SIFs were considered to have overlapping confidence intervals; consolidation was the only SIF for which the best prompt did not have overlapping confidence intervals with any of the three alternative prompts. For AVD, bronchiectasis, CAC, emphysema, and MLN, all alternative prompt versions had overlapping confidence intervals with the best prompt version. For ILA and MM, one prompt version had non-overlapping confidence intervals with the best prompt version, while for TA two out of three alternative prompt versions had non-overlapping confidence intervals with the best prompt version. For consolidation, all three alternative prompt versions had non-overlapping confidence intervals with the best prompt version. Llama-3.1-8B-INST was considered to have different agreement categorizations for four SIFs. Llama-3.1-8B-INST/Prompt V2 had substantial agreement (0.65 [0.44, 0.83]) for AVD, whilst the alternative prompts resulted in moderate or fair agreement. Llama-3.1-8B-INST/Prompt V2 had almost perfect agreement (0.83 [0.73, 0.91]) for bronchiectasis, whilst the alternative prompts resulted in substantial agreement. Llama-3.1-8B-INST/Prompt V1 had substantial agreement (0.71 [0.59, 0.83]) for consolidation, whilst the alternative prompts resulted in moderate or fair agreement. Llama-3.1-8B-INST/Prompt V1 had fair agreement (0.22 [0.16, 0.29]) for MM, while all alternative prompts had slight agreement..

**Table S2. The Fleiss Kappa for each LLM Assessing the Agreement across the Different Prompt Strategies with 95% Boostrapped Confidence Intervals**

| **IF** | **MedGemma-4B-IT** | **Llama-3.1-8B-INST** | **GPT-5** | **GPT-4-Turbo** |
| --- | --- | --- | --- | --- |
| **Aortic valve disease** | 0.60 (0.46, 0.72)  *Moderate* | 0.38 (0.24, 0.50)  *Fair* | 0.97 (0.93, 1.00)  *Almost Perfect* | 0.89 (0.78, 0.96)  *Almost Perfect* |
| **Bronchiectasis** | 0.62 (0.53, 0.70)  *Substantial* | 0.77 (0.69, 0.83)  *Substantial* | 0.80 (0.72, 0.86)  *Substantial* | 0.97 (0.93, 0.99)  *Almost Perfect* |
| **Consolidation** | 0.50 (0.42, 0.57) | 0.41 (0.32, 0.48) | 0.65 (0.56, 0.72)  *Substantial* | 0.65 (0.56, 0.72)  *Substantial* |
| **Coronary artery calcification** | 0.68 (0.62, 0.74)  *Substantial* | 0.72 (0.66, 0.78)  *Substantial* | 0.99 (0.97, 1.00)  *Almost Perfect* | 0.92 (0.89, 0.96)  *Almost Perfect* |
| **Emphysema** | 0.54 (0.46, 0.61)  *Moderate* | 0.81 (0.73, 0.87)  *Almost Perfect* | 0.99 (0.96, 1.00)  *Almost Perfect* | 0.96 (0.93, 0.99)  *Almost Perfect* |
| **Interstitial lung abnormalities** | 0.43 (0.35, 0.51)  *Moderate* | 0.59 (0.52, 0.65)  *Moderate* | 0.74 (0.67, 0.81)  *Substantial* | 0.77 (0.70, 082)  *Substantial* |
| **Mediastinal lymph nodes** | 0.72 (0.66, 0.77)  *Substantial* | 0.65 (0.58, 0.72)  *Substantial* | 0.85 (0.79, 0.89)  *Almost Perfect* | 0.85 (0.79, 0.89)  *Almost Perfect* |
| **Mediastinal mass** | 0.43 (0.36, 0.49)  *Moderate* | 0.45 (0.36, 0.54)  *Moderate* | 0.85 (0.75, 0.93)  *Almost Perfect* | 0.64 (0.52, 0.74)  *Substantial* |
| **Thyroid abnormalities** | 0.45 (0.35, 0.52)  *Moderate* | 0.46 (0.35, 0.56)  *Moderate* | 0.99 (0.97, 1.00)  *Almost Perfect* | 0.84 (076, 0.91)  *Almost Perfect* |

In Table S2 the agreement for each LLM between the four prompt versions is shown. For the open source LLMs the prompt affected the label choices more than for the proprietary LLMs. For MedGemma-4B-IT, for most of the SIFs only moderate agreement was obtained across the prompt versions, and only bronchiectasis (0.62 [0.53, 0.70]), CAC (0.68 [0.62, 0.74]), and mediastinal lymph nodes (0.72 [0.66, 0.77]) obtained substantial agreement. For Llama-3.1-8B-INST, there was more agreement amongst prompt versions, since three SIFs had substantial agreement: bronchiectasis (0.77 [0.69, 0.83]), CAC (0.72 [0.66, 0.78]), mediastinal lymph nodes (0.65 [0.58, 0.72]) and almost perfect agreement for emphysema (0.81 [0.73, 0.87]). For GPT-5, almost perfect agreement between prompt versions was obtained for seven of the SIFs, the two which had substantial agreement were consolidation (0.65 [0.56, 0.72]) and ILA (0.74 [0.67, 0.81]). For GPT-4-Turbo, almost perfect agreement was obtained for six of the SIFs, the three with only substantial agreement were: consolidation (0.65 [0.56, 0.72]), ILA (0.77, [0.70, 082]), and mediastinal mass 0.64 (0.52, 0.74).
